# The Hydrocephalus Association Patient-Powered Interactive Engagement Registry (HAPPIER): Design and Initial Baseline Report

**DOI:** 10.1101/2024.10.11.24315348

**Authors:** Noriana E. Jakopin, Samantha N. Lanjewar, Amanda Garzon, Paul Gross, Richard Holubkov, Abhay Moghekar, Jason Preston, Margaret Romanoski, Chevis N. Shannon, Mandeep S. Tamber, Tessa van der Willigen, Melissa Sloan, Monica J. Chau, Jenna E. Koschnitzky

## Abstract

**Objective:** Hydrocephalus is a neurological condition characterized by an accumulation of cerebrospinal fluid with no cure and limited treatments. There is a significant gap in hydrocephalus research where patients lack opportunities to voice their perspectives on their condition. The Hydrocephalus Association Patient-Powered Interactive Engagement Registry (HAPPIER) database was created to highlight the quality-of-life outcomes in hydrocephalus from a longitudinal perspective. HAPPIER ensures that the lived experiences of those affected by hydrocephalus are highlighted, and provides a platform for researchers to access this data or distribute their own surveys, ultimately aiming to improve patient-centered care and outcomes. This publication introduces the registry to the medical and scientific community by highlighting demographics, etiology, treatments, symptom profiles, and diagnosed comorbidities of the participants.

**Methods:** HAPPIER was developed by the Hydrocephalus Association and a 10-member steering committee. Development of its surveys was informed by other registries with similar goals, existing surveys and assessments, and input from University of Utah Data Center faculty. The study population was recruited using social and traditional media, referrals from medical professionals, and advertisement at Hydrocephalus Association-sponsored events.

**Results:** Of the 691 survey participants (referring to those with hydrocephalus), 451 (65.3%) were individuals responding for themselves. 380 (55.0%) of the registry population was female, 594 (86.0%) was white, and 606 (87.7%) was from the United States and territories. The most common age reported for diagnosis was between 0 and 11 months (46.2%), and the most common hydrocephalus etiology reported was congenital hydrocephalus (43.8%). The most prevalent treatment reported was a shunt(s) (71.2%). The most commonly-reported symptom was headaches (60.3%), and 69.9% of participants reported being diagnosed with movement impairments and 70.8% with other health conditions.

**Conclusion:** HAPPIER is a novel database developed to address the gaps in data in non-clinical outcomes of hydrocephalus, which are critical to clinical care and understanding hydrocephalus in its totality. Patient perspectives and outcomes have been historically underrepresented. By directly engaging individuals living with hydrocephalus and their caregivers, HAPPIER is designed to incorporate essential patient perspectives through planned longitudinal data collection and patient surveys. These data are open to investigators interested in analyzing the collected data.

## INTRODUCTION

Hydrocephalus is a chronic, multifaceted condition with no cure characterized by an abnormal accumulation of cerebrospinal fluid (CSF) in the ventricles of the brain. Many individuals with hydrocephalus experience a variety of motor, cognitive, neuropsychiatric, quality of life, and developmental impairments^1–8^. Hydrocephalus treatments are limited to a surgically inserted ventricular or lumbar shunt, or an endoscopic third ventriculostomy (ETV) with or without choroid plexus cauterization (CPC), all of which have high rates of failure^9^. Major gaps exist in hydrocephalus research when it comes to tracking, understanding, diagnosing, and ameliorating these outcomes. Much of the existing data comes from clinical research networks, notably the Hydrocephalus Clinical Research Network (HCRN)^10,11^ and the Adult Hydrocephalus Research Network (AHCRN)^12^. Surgical and clinical data are available, but with limited information from the patient perspective. Despite the chronic nature of hydrocephalus, there is limited infrastructure for the collection of longitudinal data, especially for the quality of life from the patient perspective.

In response to these gaps, the Hydrocephalus Association developed the Hydrocephalus Association Patient-Powered Interactive Engagement Registry (HAPPIER). This survey-based patient registry is longitudinal and allows for collection of a wide range of data relating to the life course and experiences of those with hydrocephalus. HAPPIER was conceived with four primary goals in mind: (1) to provide investigators and clinicians with disease symptoms from a patient perspective, current treatment practices, and patient-centered outcomes to better guide therapeutic interventions; (2) to provide longitudinal descriptive data to generate project hypotheses by the Hydrocephalus Association and outside researchers; (3) to track the characteristics and experiences of those living with hydrocephalus over time; and (4) to identify patients potentially eligible for research studies and/or clinical trials. The registry also provides a readily accessible population for other survey-based studies to be developed. This article will report how HAPPIER was built to address the data gaps in the field and provide the initial findings on the demographics and characteristics of the registry population.

## METHODS

### Design

A 10-member steering committee composed of Hydrocephalus Association staff, scientists and medical professionals in the field of hydrocephalus, persons living with hydrocephalus, and caregivers of those with hydrocephalus was formed in 2015. Registries with similar goals, such as the Interactive Autism Network^13^ and Cystic Fibrosis Foundation Patient Registry Data^14^, were studied along with other surveys and assessments, including the NINDS Quality of Life in Neurological Disorders (Neuro-QoL)^15^, NIH Patient-Reported Outcomes Measurement Information System (PROMIS)^16^, 15D Quality of Life Questionnaire^17^, National Alzheimer’s Coordinating Center (NACC) Functional Activities Questionnaire (FAQ)^18^, Katz Index of Independence in Activities of Daily Living^19,20^, Lawton-Brody Instrumental Activities of Daily Living Scale (I.A.D.L.)^21^, Pediatric Quality of Life Inventory^22^, and Hydrocephalus Outcome Questionnaire^23^. The faculty at the University of Utah Data Center and other experts also provided input. These sources informed the development of HAPPIER. Care was taken to ensure that the survey was understandable.

The web-based Hydrocephalus Association Patient Portal was developed by the Hydrocephalus Association for participants to access and enter the registry. Data from participants over the age of 18 and from caregivers were collected and managed through a REDCap database instance hosted at the University of Utah. REDCap is a platform designed for data capture, management, and collection pertaining to surveys, databases, trials, applications, questionnaires, registries, quality reviews, and various types of research studies^24^.

### Ethics Statement

Approval was obtained from the University of Utah Institutional Review Board (IRB) in May 2017. The study was approved for exemption status because it poses minimal risk, is in accordance with the Belmont report, and contains orderly monitoring and accounting of research activities. Upon entry into the portal, registrants accessed a consent letter providing them with information about filling out the surveys and with contact information of the study investigators. Only patients over the age of 18 years old with the ability to provide consent took the survey for themselves. For patients under the age of 18 years old, or patients who had impaired ability to provide consent, caregivers filled out the surveys in their stead. All data were stored in REDCap. The server facility was located separately from the remainder of the Data Center at the University of Utah. All datasets used for analysis were fully de-identified.

### Recruitment

Social and traditional media were developed for participant recruitment through the Hydrocephalus Association, including emails, posts on Facebook, Twitter, and Instagram, printed newsletters, direct mail, and advertisement on the Hydrocephalus Association’s website. Medical professionals were also asked to refer patients to join HAPPIER, and HAPPIER was advertised at Hydrocephalus Association-sponsored events, such as community-centered conferences and fundraising events. All potential participants were pointed to the Hydrocephalus Association Patient Portal for more information and instructions to join the registry.

### Analysis

HAPPIER was activated on June 29^th^, 2018. The dataset for this analysis consisted of responses to the baseline survey (Supplemental File 1) and was retrieved on March 8^th^, 2022. The registry population’s demographic makeup, hydrocephalus etiology breakdown, and treatments received were recorded. Comorbidities, symptom categories, and treatment history were captured from survey responses and reported as percentages of the overall registry population. A broad view of the presence of comorbidities and symptom categories was also captured from survey responses. This data was reported as percentages of the registry population who experienced various comorbidities and symptoms. Treatment history was also reported in the same manner. The percentage of the population who underwent shunt and/or ETV revision surgery and who experienced shunt infection was determined. Finally, the retention rate for the longitudinal arm of the registry was calculated.

### Data Access Statement

To obtain data from the HAPPIER registry to conduct a study, or to use HAPPIER to distribute a new survey, please email the Hydrocephalus Association National Director of Research.

## RESULTS

### Initial Population Characteristics

The total number of people that participated in the HAPPIER registry and completed the baseline survey (Supplemental File 1) was 691. Participants refer to the individuals with hydrocephalus represented in the survey data, both those who recorded their own responses and those for whom a caregiver answered on their behalf. The HAPPIER registry captures demographic information for all participants (Table 1). 65.3% of the participants were persons living with hydrocephalus and completed the surveys for themselves, and 34.7% had a caregiver complete the surveys in their stead. At the time of the survey, 72.2% of participants were over the age of 18. Data was represented from all ages including infants (0-11 months, 5.8%), toddlers (1-3 years, 9.1%), children (4-12 years, 9.0%), teenagers (13-17 years, 3.9%), young adults (18-34 years, 32.1%), adults (35-59 years, 27.4%), and older adults (60+ years, 12.7%). There was a majority of female participants (55.0%), and males represented 44.3%. Other respondents, who either preferred not to answer or identified as transsexual, accounted for 0.7% of the total participants.

**Table 1.**
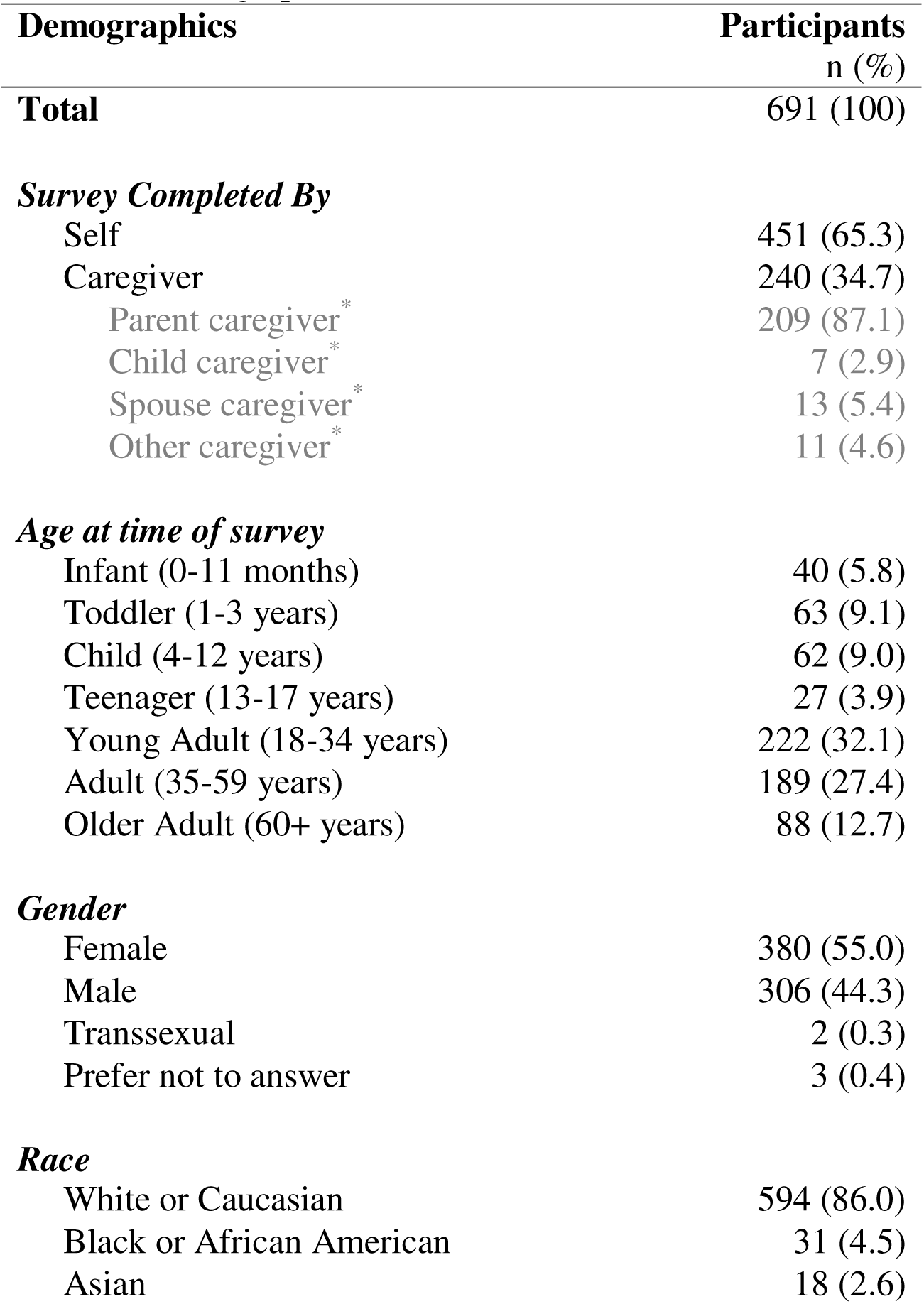
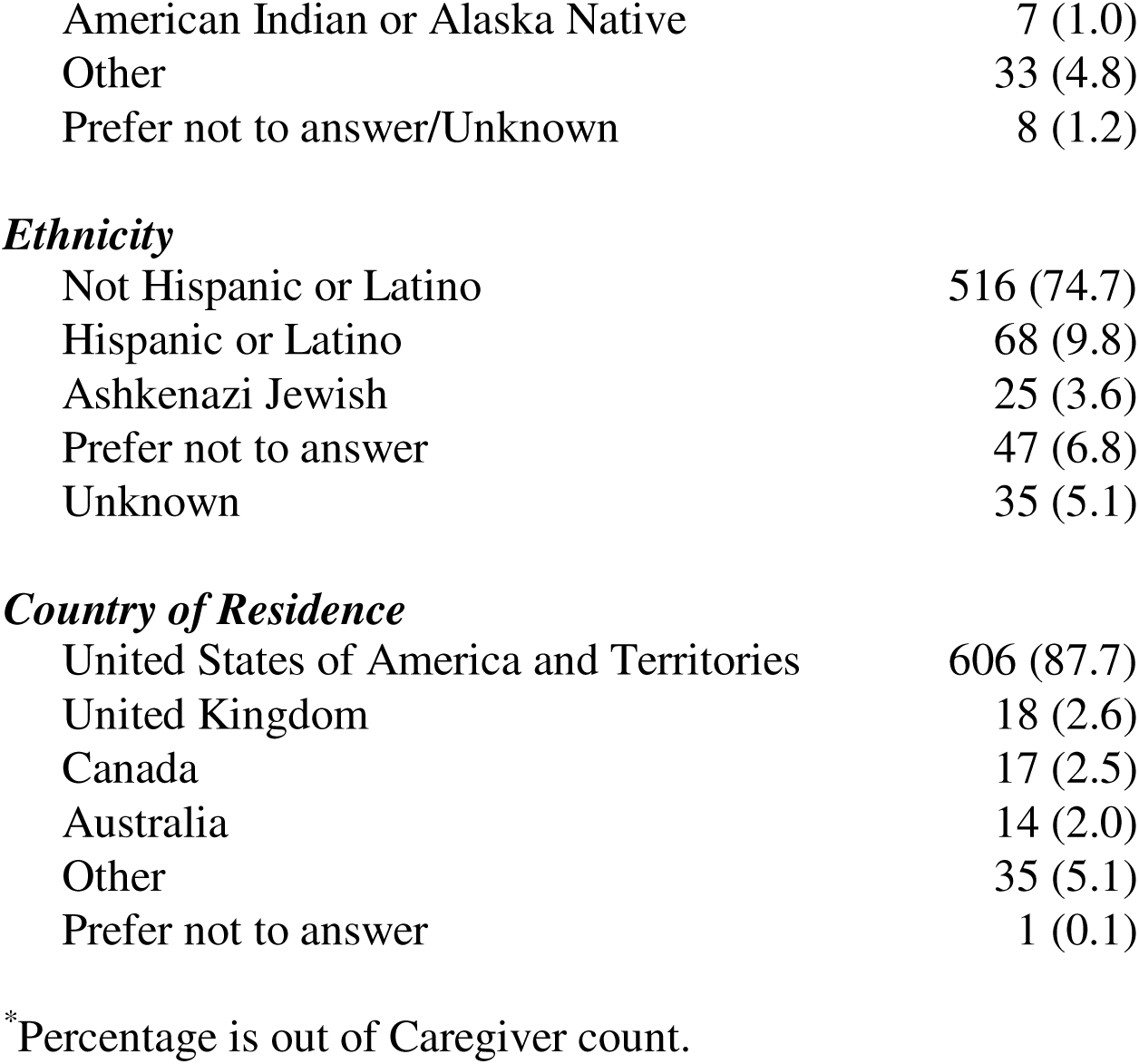
Demographics.

| **Demographics** | **Participants**<br>**n (%)** |
| --- | --- |
| **Total** | 691 (100) |
| ***Survey Completed By*** |  |
| Self | 451 (65.3) |
| Caregiver | 240 (34.7) |
| Parent caregiver* | 209 (87.1) |
| Child caregiver* | 7 (2.9) |
| Spouse caregiver* | 13 (5.4) |
| Other caregiver* | 11 (4.6) |
| ***Age at time of survey*** |  |
| Infant (0-11 months) | 40 (5.8) |
| Toddler (1-3 years) | 63 (9.1) |
| Child (4-12 years) | 62 (9.0) |
| Teenager (13-17 years) | 27 (3.9) |
| Young Adult (18-34 years) | 222 (32.1) |
| Adult (35-59 years) | 189 (27.4) |
| Older Adult (60+ years) | 88 (12.7) |
| ***Gender*** |  |
| Female | 380 (55.0) |
| Male | 306 (44.3) |
| Transsexual | 2 (0.3) |
| Prefer not to answer | 3 (0.4) |
| ***Race*** |  |
| White or Caucasian | 594 (86.0) |
| Black or African American | 31 (4.5) |
| Asian | 18 (2.6) |
| American Indian or Alaska Native | 7 (1.0) |
| Other | 33 (4.8) |
| Prefer not to answer/Unknown | 8 (1.2) |
| ***Ethnicity*** |  |
| Not Hispanic or Latino | 516 (74.7) |
| Hispanic or Latino | 68 (9.8) |
| Ashkenazi Jewish | 25 (3.6) |
| Prefer not to answer | 47 (6.8) |
| Unknown | 35 (5.1) |
| ***Country of Residence*** |  |
| United States of America and Territories | 606 (87.7) |
| United Kingdom | 18 (2.6) |
| Canada | 17 (2.5) |
| Australia | 14 (2.0) |
| Other | 35 (5.1) |
| Prefer not to answer | 1 (0.1) |

\*Percentage is out of Caregiver count.

The participants predominantly identified their race as white (86.0%). Black or African American participants accounted for 4.5% of the participants, Asian, 2.6%, American Indian or Alaska Native, 1.0%. Those who identified in the “other” group accounted for 4.8% of participants, and those who marked “preferred not to answer or unknown” accounted for 1.2% of participants. The participants indicated their ethnicities as follows: not Hispanic or Latino (74.7%), Hispanic or Latino (9.8%), Ashkenazi Jewish (3.6%), preferred not to answer (6.8%), or selected “unknown” (5.1%). Most participants resided in the United States or its territories (87.7%). Participants living in the United Kingdom, Canada, and Australia made up 7.1% of the registry. The remaining participants were from countries including Argentina, Bahamas, Belgium, Denmark, Egypt, France, Germany, Hungary, India, Ireland, Israel, Jordan, Kenya, Nepal, Netherlands, New Zealand, Norway, Philippines, Portugal, Russia, South Africa, Turkey, Uruguay, and Zimbabwe. One person preferred not to answer (0.1%).

The survey captured participants’ hydrocephalus-specific characteristics (Table 2 and Figure 1). Most participants were diagnosed before the age of one, as infants (0-11 months) or prenatally (58.9%). Others were diagnosed in the following years: toddler (1-3 years, 4.1%), child (4-12 years, 5.6%), teenager (13-17 years, 3.6%), young adult (18-34 years, 8.5%), adult

**Figure 1.**
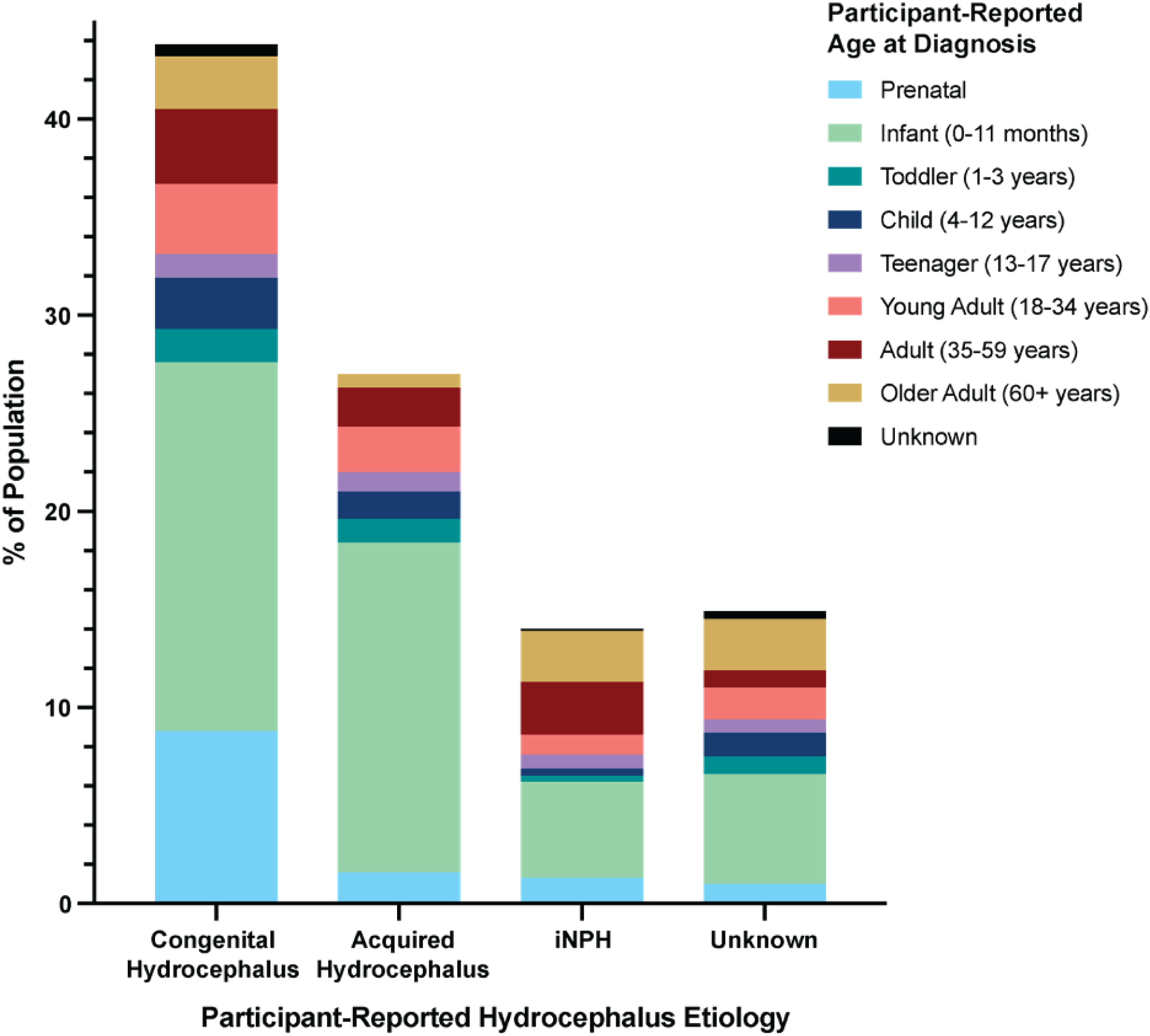
Participant-Reported Hydrocephalus Etiology and Age at Diagnosis. Data are self-reported from patients or their caregivers, and not based on data from medical professionals. The age of diagnosis breakdown for congenital hydrocephalus, acquired hydrocephalus, idiopathic normal pressure hydrocephalus (iNPH), and unknown/unsure or prefer not to answer etiologies is displayed. The highest number of participants (43.8%) had congenital hydrocephalus, with 42.3% of this group reported being infants at the age of diagnosis.

**Table 2.**
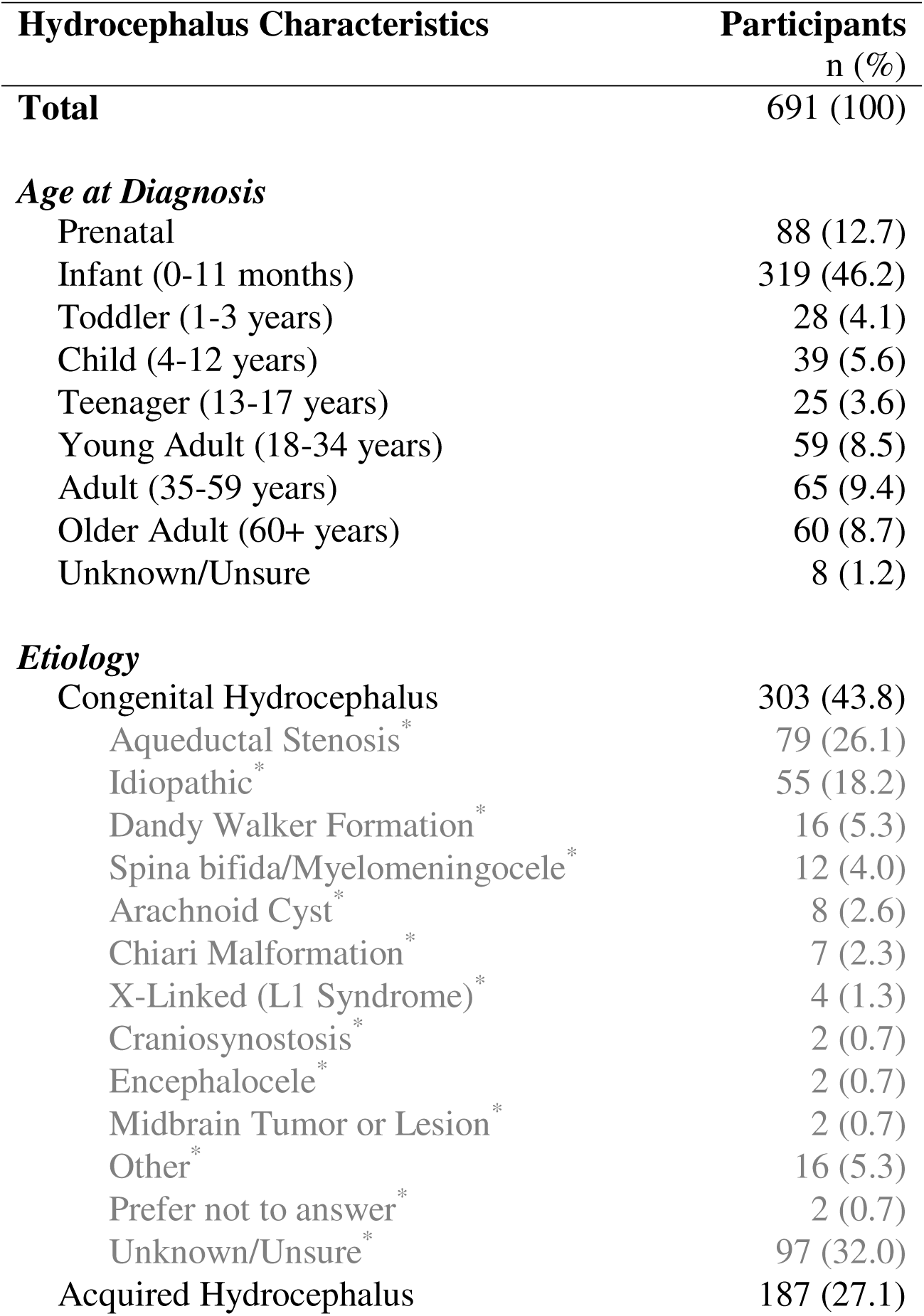
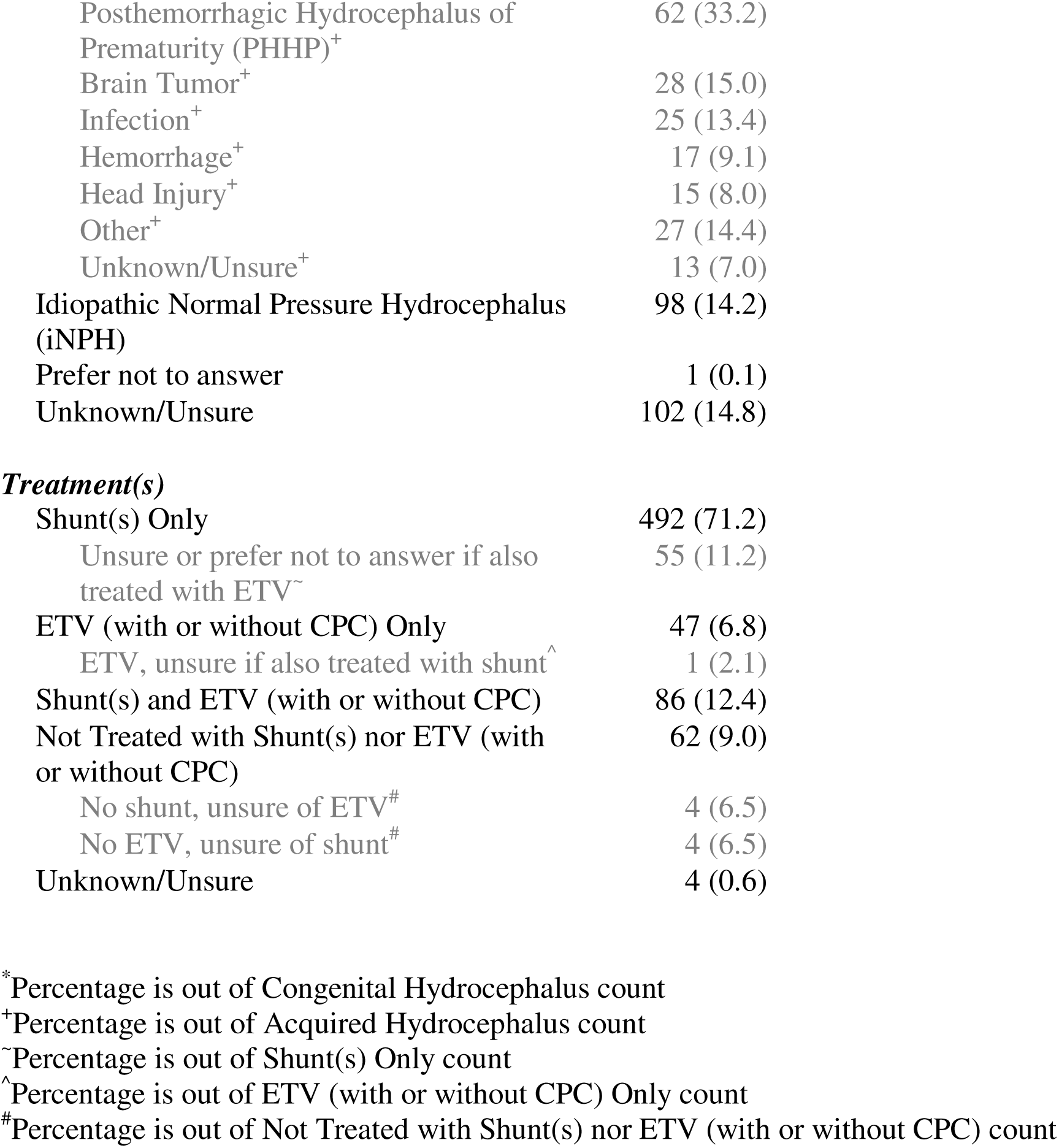
Hydrocephalus Characteristics.

| **Hydrocephalus Characteristics** | **Participants**<br>**n (%)** |
| --- | --- |
| **Total** | 691 (100) |
| ***Age at Diagnosis*** |  |
| Prenatal | 88 (12.7) |
| Infant (0-11 months) | 319 (46.2) |
| Toddler (1-3 years) | 28 (4.1) |
| Child (4-12 years) | 39 (5.6) |
| Teenager (13-17 years) | 25 (3.6) |
| Young Adult (18-34 years) | 59 (8.5) |
| Adult (35-59 years) | 65 (9.4) |
| Older Adult (60+ years) | 60 (8.7) |
| Unknown/Unsure | 8 (1.2) |
| ***Etiology*** |  |
| Congenital Hydrocephalus | 303 (43.8) |
| Aqueductal Stenosis * | 79 (26.1) |
| Idiopathic * | 55 (18.2) |
| Dandy Walker Formation * | 16 (5.3) |
| Spina bifida/Myelomeningocele * | 12 (4.0) |
| Arachnoid Cyst * | 8 (2.6) |
| Chiari Malformation * | 7 (2.3) |
| X-Linked (L1 Syndrome) * | 4 (1.3) |
| Craniosynostosis * | 2 (0.7) |
| Encephalocele * | 2 (0.7) |
| Midbrain Tumor or Lesion * | 2 (0.7) |
| Other * | 16 (5.3) |
| Prefer not to answer * | 2 (0.7) |
| Unknown/Unsure * | 97 (32.0) |
| Acquired Hydrocephalus | 187 (27.1) |
| Posthemorrhagic Hydrocephalus of Prematurity (PHHP)<sup>+</sup> | 62 (33.2) |
| Brain Tumor<sup>+</sup> | 28 (15.0) |
| Infection<sup>+</sup> | 25 (13.4) |
| Hemorrhage<sup>+</sup> | 17 (9.1) |
| Head Injury<sup>+</sup> | 15 (8.0) |
| Other<sup>+</sup> | 27 (14.4) |
| Unknown/Unsure<sup>+</sup> | 13 (7.0) |
| Idiopathic Normal Pressure Hydrocephalus (iNPH) | 98 (14.2) |
| Prefer not to answer | 1 (0.1) |
| Unknown/Unsure | 102 (14.8) |
| ***Treatment(s)*** |  |
| Shunt(s) Only | 492 (71.2) |
| Unsure or prefer not to answer if also treated with ETV<sup>~</sup> | 55 (11.2) |
| ETV (with or without CPC) Only | 47 (6.8) |
| ETV, unsure if also treated with shunt<sup>^</sup> | 1 (2.1) |
| Shunt(s) and ETV (with or without CPC) | 86 (12.4) |
| Not Treated with Shunt(s) nor ETV (with or without CPC) | 62 (9.0) |
| No shunt, unsure of ETV<sup>#</sup> | 4 (6.5) |
| No ETV, unsure of shunt<sup>#</sup> | 4 (6.5) |
| Unknown/Unsure | 4 (0.6) |

\*Percentage is out of Congenital Hydrocephalus count
<sup>+</sup>Percentage is out of Acquired Hydrocephalus count
<sup>~</sup>Percentage is out of Shunt(s) Only count
<sup>^</sup>Percentage is out of ETV (with or without CPC) Only count
<sup>#</sup>Percentage is out of Not Treated with Shunt(s) nor ETV (with or without CPC) count

(35-59 years, 9.4%), older Adult (60+ years, 8.7%), and 1.2% answered “unknown/unsure”. Participants reported many types of causes of hydrocephalus, which were divided into the major categories of congenital hydrocephalus, acquired hydrocephalus, idiopathic normal pressure hydrocephalus (iNPH), unknown/unsure, and prefer not to answer (Table 2). Congenital hydrocephalus was the most prevalent cause of hydrocephalus amongst participants (43.8%), followed by acquired hydrocephalus (27.1%). The top reported subcategories of congenital hydrocephalus included those who reported unknown/unsure (32%), aqueductal stenosis (26.1%), and idiopathic hydrocephalus (18.2%). The top reported subcategories of acquired hydrocephalus included posthemorrhagic hydrocephalus of prematurity (PHHP, 33.2%), brain tumor (15.0%), and infection (13.4%). 14.2% of participants reported being diagnosed with idiopathic normal pressure hydrocephalus (iNPH).

The most prevalent treatment was with a shunt (Table 2). There were 492 (71.2%) participants who were treated with shunt(s) only. Of these 492 participants, 55 (11.2%) were unsure or preferred not to answer if they had also been treated with an endoscopic third ventriculostomy (ETV). Other participants reported only being treated with an ETV (with or without CPC) (6.8%), with one person (2.1%) uncertain if they had also been treated with a shunt. Participants also reported being treated with shunt(s) and ETV (with or without CPC) (12.4%), neither treatment (62 participants, 9.0%), or were unsure or did not know of their treatment (0.6%). Of the 62 participants who had not been treated with shunt(s) nor ETV (with or without CPC), 4 participants (6.5%) had no shunt but were unsure if they were treated with ETV, and 4 participants (6.5%) had no ETV but were unsure if they were treated with a shunt.

Participants also reported their treatment complications (Figure 2). Of the 578 participants who were treated with a shunt(s) (including those with shunt(s) and ETV), 399 (69.0%) of them had at least one shunt revision, and 138 (23.9%) of them had at least one shunt infection. Of the 133 participants who were treated with an ETV (with or without CPC) (also including those with shunt(s) and ETV), 46 (34.6%) of them reported receiving at least one ETV revision. Among those who experienced an ETV revision, 69.6% had 1 revision, 23.9% had 2-4 revisions, and 6.5% had 5 or more revisions.

**Figure 2.**
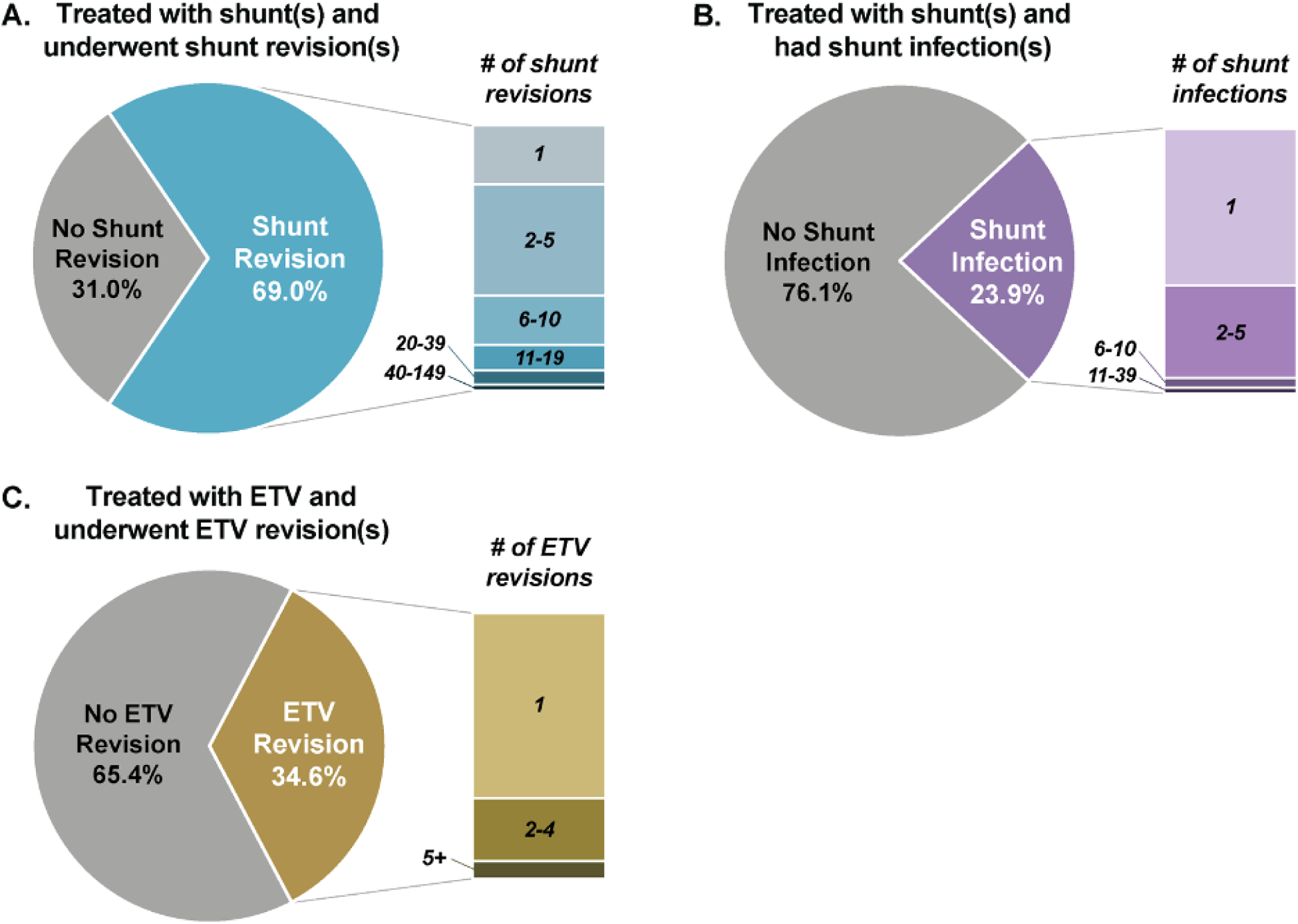
Treatment Complications. Displays the percentage of participants treated with shunt(s) and (A) underwent shunt revision(s) or (B) had shunt infection(s), and the percentage of participants treated with ETV that (C) underwent ETV revision(s), along with the number of shunt revisions, shunt infections, and ETV revisions participants have had, respectively. Those treated with shunt(s) includes all participants treated with shunt(s) only or shunt(s) and ETV (with or without CPC). Those treated with ETV includes all participants treated with ETV (with or without CPC) only or shunt(s) and ETV (with or without CPC). Shunt revisions were the most common treatment complication.

Participants reported their symptoms (Figure 3). The three most frequently reported symptoms were headaches (60.3%), tiredness/lethargy (55.4%), and memory problems (47.6%). Participants also reported their diagnosed comorbidities, which were grouped into the following general categories: movement impairments, emotional disorders, cognitive disorders, educational disorders, and other diagnosed health conditions (Figure 4). 69.9% of the survey population had reported movement impairments including tremors, ticks, and other stereotyped movements, or difficulty with balance and walking. Emotional disorders, including depression, anxiety, and/or OCD, were diagnosed in 44.7% of registrants. Cognitive disorders, including ADHD, autism, receptive delay, expressive delay, executive function disorder, global developmental delay, intellectual disability, visual processing deficits, and short-term memory problems, were diagnosed in 42.5% of the registrants. Educational disorders, including learning disabilities in math, reading, and/or writing and non-verbal learning disabilities, were diagnosed in 26.6% of registrants. 70.8% received a diagnosis for at least one other diagnosed health condition, aside from hydrocephalus, with the top three most prevalent conditions being visual impairment, migraine, and chronic pain. Other conditions within this category include epilepsy, cerebral palsy, Chiari malformation, spina bifida, sleep apnea, diabetes, obesity, hypertension, and various dementias, amongst others.

**Figure 3.**
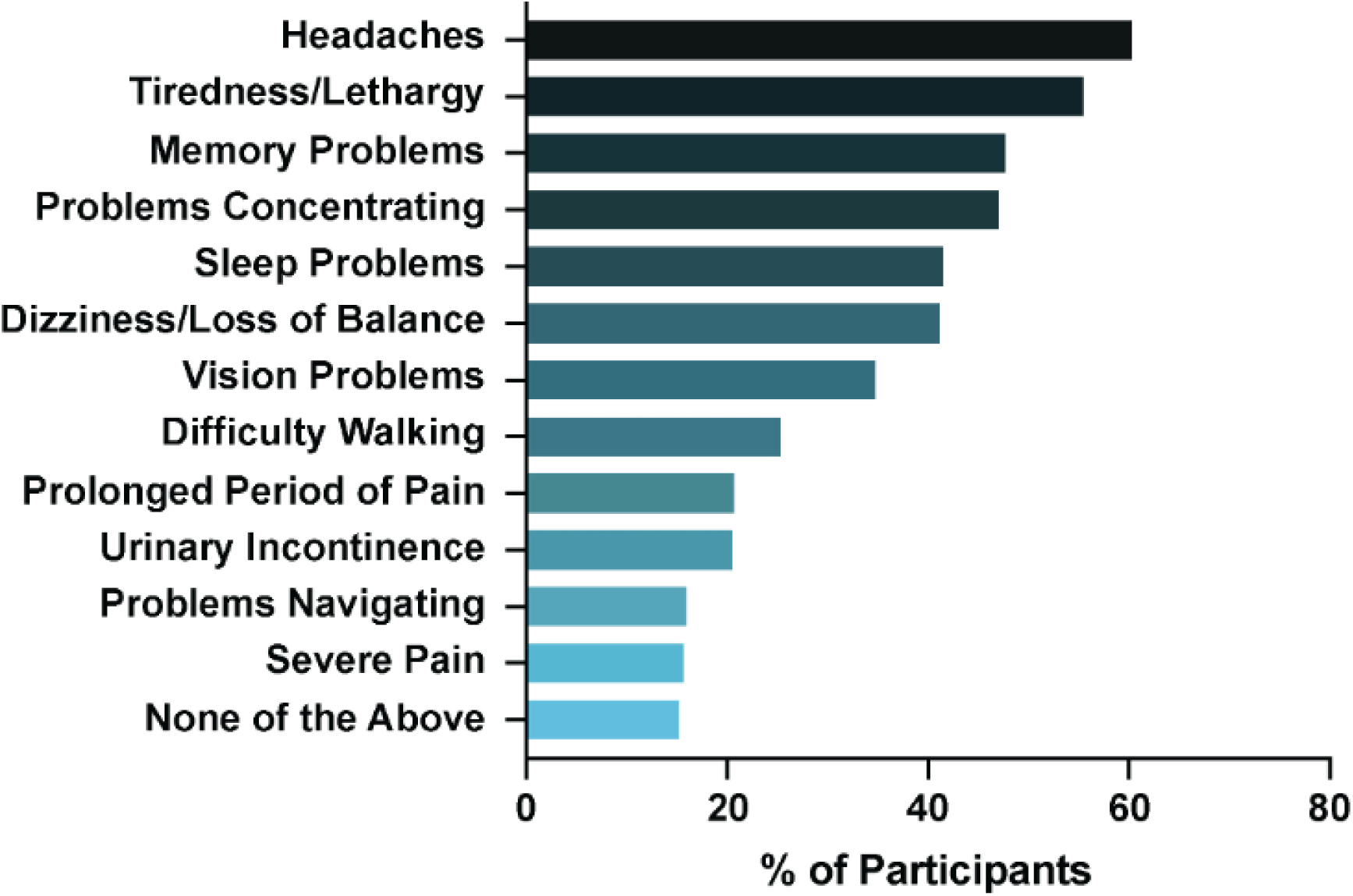
Symptoms Profile. Displays the percentage of participants experiencing symptoms most associated with hydrocephalus. Headaches, tiredness/lethargy, and memory problems were the most frequently reported symptoms.

**Figure 4.**
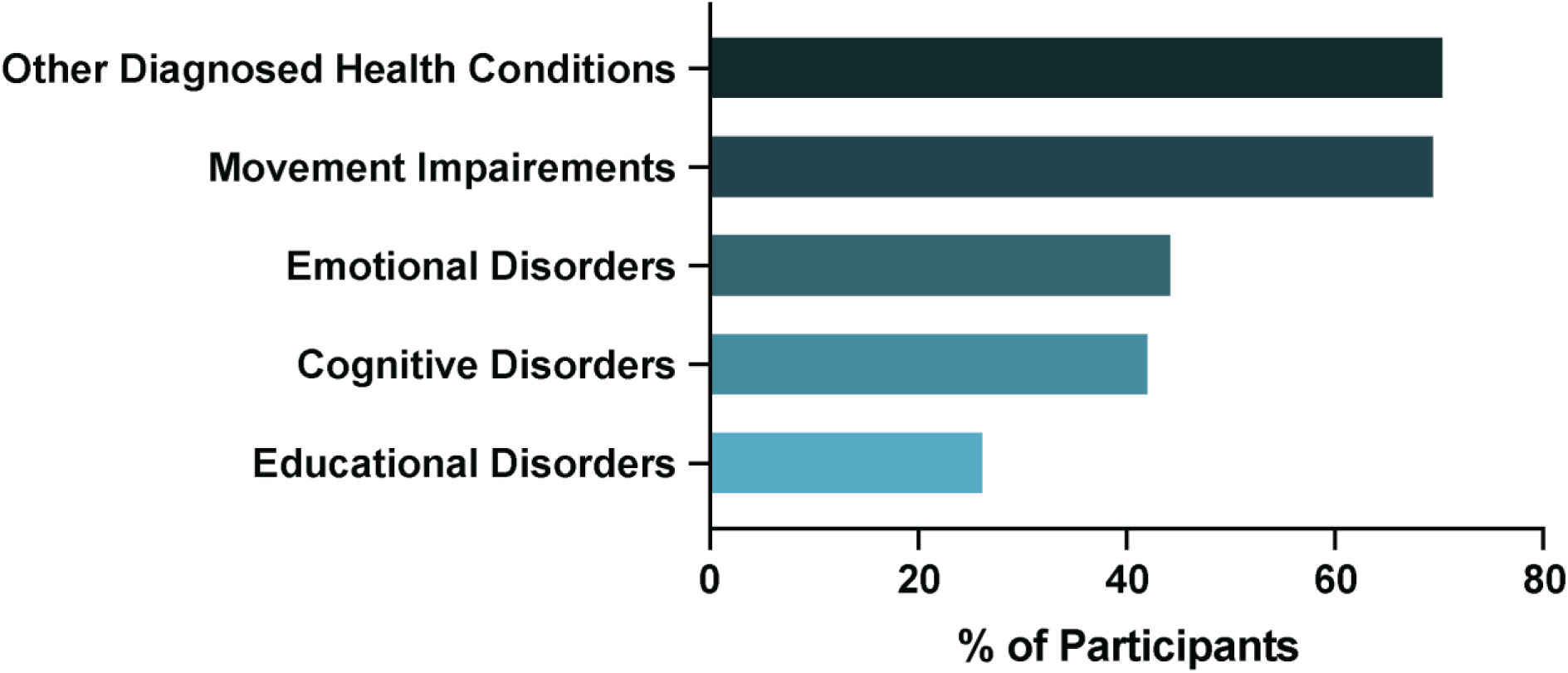
Diagnosed Comorbidities. Displays the percentage of participants experiencing other diagnosed health conditions, impairments, or disorders alongside hydrocephalus.

One of the goals of HAPPIER is to collect longitudinal data. Between 2018 and 2020, 618 participants joined HAPPIER and completed the baseline survey (Supplemental File 1). Of this population, 35% continued with the longitudinal arms and completed the 2021 Annual Survey.

## DISCUSSION

HAPPIER was created to capture the perspectives, experiences and outcomes of patients with hydrocephalus to address a significant gap in patient-centered data in hydrocephalus research. The registry infrastructure is built to collect longitudinal data on measures not typically found in medical records, such as quality of life measures, as these outcomes have been historically lacking. The purpose of this initial publication is to present an overview of the composition of the registry, highlighting the demographics of the participants, the etiology of hydrocephalus, treatments, complications, symptom profiles, and comorbidities.

The initial participation from 691 people indicates that the hydrocephalus patient community wants to share their information with the medical and scientific communities. The data captured from all different age ranges, from newborns through older adults (up to 90 years old), emphasizes the desire for a wide range of patients and caregivers to participate in research and contribute their perspective. The racial demographics of the HAPPIER registry, with 86.0% of participants identifying as white, present a discrepancy compared to observed national hydrocephalus data^25–27^ which demonstrate a more diverse patient population. This underrepresentation underscores the need for targeted efforts to increase diversity within the HAPPIER dataset to accurately reflect the broader hydrocephalus community and further support equitable research and care. Furthermore, when the registry was initially created in 2018, the gender options were limited to “male”, “female”, “transexual”, and “prefer not to answer”. At the time of data retrieval, 2 participants identified as transexual and 3 selected prefer not to answer. Given the restricted choices, it is possible that participants may have responded differently had more inclusive options, such as those now available, been provided in the original survey. The Hydrocephalus Association has since updated the HAPPIER survey options for gender identity, modeled after the NIH’s All of Us Program categories, to include “male”, “female”, “non-binary”, “transgender”, “prefer not to answer”, and an area to list their own description.

In addition to capturing demographic information, an important aspect of the registry is that it sheds light on the multifaceted health challenges faced by participants, some of which are not typically captured during medical visits. These patient-reported symptoms and comorbidities highlight the complex experiences of those living with hydrocephalus. This suggests that available treatments do not adequately address motor, cognitive, developmental, and neuropsychiatric difficulties present with hydrocephalus. Furthermore, seeing as 9.0% of registrants are not treated with a shunt nor an ETV, it is worth investigating this subset of the population further to see if these individuals represent a group that needs treatment but is not being treated.

A vast majority of the surveyed population had at least one other diagnosed health condition in addition to hydrocephalus (70.8%). Among those living with iNPH, those with multiple comorbidities are reported to have worse hydrocephalus symptoms prior to any operation, especially regarding gait and balance performance^28^. These comorbidities can also worsen the prognosis of the condition and of CSF shunting^29^. Patients with comorbidities may require a different approach to treatment than those without^29^, and HAPPIER results suggest that the percentage of patients falling in this category is substantial. Furthermore, by reporting data on which symptoms are present among the highest percentage of registrants, as shown in Figure 3, HAPPIER captures information that could be beneficial in future research to determine potential new treatments to target the most prevalent symptoms.

The etiology breakdown among HAPPIER participants provides insights alongside the current literature. In a survey that collected data from 683 children and adults with hydrocephalus in a neurosurgical center over five years, it was found that 30.8% had hydrocephalus that stemmed from a congenital malformation and 8.9% had NPH^30^. In contrast, a study that extracted hospital admissions data from the Healthcare Cost and Utilization Project (HCUP), National Inpatient Sample (NIS), Agency for Healthcare Research and Quality (AHRQ), and Kids’ Inpatient Database (KID) reported that 8.0% of hydrocephalus etiology hospital admissions were for congenital hydrocephalus^26^. Another study of 1,000 individuals aged 65+ in Sweden reported a prevalence rate of 3.7% for iNPH^31^. Discordantly, our data reported a higher figure for both congenital hydrocephalus (43.8%) and iNPH (14.2%). These varying numbers may be a reflection of our recruitment methods, which included targeting of community network support groups, potentially skewing the data towards these populations. This indicates a need for increased recruitment efforts and diversity within the registry to ensure all etiologies are properly represented and reflective of the national averages.

Similarly, these disparities may be due to the self-reporting nature of the patient registry and potential misinformation about what type of hydrocephalus the participant has. Since HAPPIER is not based on clinical data from medical professionals, the data reveals that a considerable number of hydrocephalus caregivers and patients have substantial knowledge gaps and uncertainty, particularly regarding hydrocephalus etiology. Notably, 14.8% of HAPPIER participants did not know or were not sure of the type of hydrocephalus they had. Furthermore, 32.0% of those with congenital hydrocephalus and 7.0% of those with acquired hydrocephalus were unaware of the cause of their condition. Surprisingly, the vast majority (80.6%) of participants who reported an iNPH diagnosis recorded that they were diagnosed under the age of 60 (*e.g.* as infants or young adults), and 18.4% over the age of 60 (Figure 1). This reveals that participants had a great misunderstanding of what iNPH is. iNPH primarily affects those 65 and older^32^. Their misunderstanding may have stemmed from their interpretation that if their hydrocephalus is well-controlled and “normal”, they have “normal” pressure hydrocephalus. Taken together, this data reveals that a substantial number of participants may lack knowledge about their overall medical profile, which could lead to more difficulty accessing treatment and advocating for themselves. This demonstrates a great need for better patient education, underscoring the value of patient-reported registries, which can capture insights directly from patients and their caregivers, addressing gaps in understanding and highlighting unmet needs^32^. One potential solution to this lack of patient knowledge is HydroAssist, an app designed to help individuals track their hydrocephalus history and better manage their condition (www.HydroAssoc.org/HydroAssist).

Identifying the unmet needs of patients can help guide changes in both clinical practice and research focus. For example, the Hydrocephalus Association conducted a formalized survey study, collecting responses from patients and family members, as well as scientists and physicians working in hydrocephalus, to determine key priorities in hydrocephalus research^33^. These responses directly informed the creation of the Hydrocephalus Association’s community research priorities^33^, which now guide their research initiatives and strategic plans. This highlights the critical role that patient-reported data plays in driving meaningful advancements in research, ensuring that efforts align with the real-world needs and concerns of the hydrocephalus community.

The HAPPIER registry offers a unique view of the population of people living with hydrocephalus, however there are some limitations of the data. There is likely a bias toward those more heavily impacted by the condition as that population is more engaged with the Hydrocephalus Association, who handled most of the recruitment. The Hydrocephalus Association advertised for patients or their caregivers to sign up for the registry and take the surveys. Future studies conducted using HAPPIER data will consider how to implement targeted recruitment of the groups missing in this initial population in terms of disease impact, as well as demographic factors, such as race and ethnicity. Additionally, as either caregivers or patients can enter the registry, there is a risk of duplicative data if both enter the registry. Software and data strategies to reduce the risk of analyzing duplicative data are being developed as well as studies to compare the matched caregiver and patient data.

This registry was created to collect longitudinal data. A 35% retention rate of engagement in the longitudinal arms indicates that a notable percentage of the initial population demonstrated an interest in continuing to engage in the registry and provide valuable data for longitudinal studies. This noted, the Hydrocephalus Association is creating a marketing and communication strategy to encourage annual compliance after the initial entry to the registry and ensure that this retention rate does not decrease over time.

Enrollment to the registry is currently active as well as data collection from these initial participants. Given the recruitment efforts to the registry and the information captured, HAPPIER has great potential as a platform for other scientists to run their own surveys and studies on this registry population. The datasets provided by HAPPIER offer unique insight into those living with hydrocephalus and allow for future investigations to better understand the condition. It has potential to identify relevant patient-reported outcomes for prospective clinical trials going forward. It can also be used in conjunction with other databases or registries, as well as by academic and private institutions, to combine or compare the data acquired with other populations. Data scientists and other hydrocephalus researchers may particularly benefit from comparing HAPPIER results with results from registries that are not self-reported to see how bias may have affected responses. Overall, HAPPIER serves as a data registry for hydrocephalus from the patient-perspective, particularly in the United States, as no other registry serves the same purpose.

## CONCLUSION

HAPPIER is a novel tool developed by the Hydrocephalus Association to address the gaps in data and infrastructure on non-clinical outcomes, which are critical to clinical care and to understanding hydrocephalus in its totality. HAPPIER collects information regarding demographics, treatment, medical history, education, and lifestyle, and tracks the changes in these variables annually. Longitudinal studies like these are important to see how reported symptoms and comorbidities, and the shunt and ETV revision frequencies may change over time and with aging. While limitations exist due to its self-reported nature, that methodology allows for this broad scope. The registry establishes a method and a population for researchers to conduct studies of various types by using the currently collected outcomes, distributing newly developed surveys, and potentially connecting clinical trials and eligible patients.

## Data Availability Statement

This data is accessible by emailing the Director of Research at the Hydrocephalus Association

