## Supplemental File 1 for "The Hydrocephalus Association Patient-Powered Interactive Engagement Registry (HAPPIER): Design and Initial Baseline Report"

### HAPPIER: Baseline Survey

Please complete the survey below.

Thank you!

**DEMOGRAPHIC DATA**
**Please fill in the information below for [initial_arm_1][name_patient]. This information will help us identify the surveys and research studies that [initial_arm_1][name_patient] is eligible for.**

**PLEASE NOTE: Once you have completed a survey you cannot make changes to that survey. If in doubt, you can save the survey at any point and return to it once you have clarified information, e.g. with your doctor.**

[initial_arm_1][name_patient]'s Date of Birth (DOB): ________________________________

Age (in years) ________________________________

[initial_arm_1][name_patient]'s gender:

- ◯ Male
- ◯ Female
- ◯ Transsexual
- ◯ Other
- ◯ Unknown
- ◯ Prefer not to answer

[initial_arm_1][name_patient]'s Race:

- ◯ American Indian or Alaska Native
- ◯ Asian
- ◯ Black or African-American
- ◯ Native Hawaiian or Other Pacific Islander
- ◯ White
- ◯ Other
- ◯ Unknown
- ◯ Prefer not to answer

[initial_arm_1][name_patient]'s ethnicity:

- ◯ Hispanic or Latino
- ◯ Not Hispanic or Latino
- ◯ Ashkenazi Jewish
- ◯ Unknown
- ◯ Prefer not to answer

#### Current residence of [initial_arm_1][name_patient]:

Country:

◯ USA
◯ Canada
◯ Afghanistan
◯ Algeria
◯ Argentina
◯ Australia
◯ Austria
◯ Bahamas
◯ Bangladesh
◯ Belgium
◯ Belize
◯ Bhutan
◯ Bolivia
◯ Bosnia and Herzegoniva
◯ Brazil
◯ Brunei
◯ Cambodia
◯ Chile
◯ China
◯ Colombia
◯ Costa Rica
◯ Cuba
◯ Cyprus
◯ Czech Republic
◯ Denmark
◯ Dominican Republic
◯ Ecuador
◯ Egypt
◯ El Salvador
◯ Ethiopia
◯ Finland
◯ France
◯ Germany
◯ Ghana
◯ Greece
◯ Guatemala
◯ Guinea
◯ Haiti
◯ Hong Kong
◯ Hungary
◯ Iceland
◯ India
◯ Indonesia
◯ Iran
◯ Iraq
◯ Ireland
◯ Israel
◯ Italy
◯ Jamaica
◯ Japan
◯ Jordan
◯ Kenya
◯ Kuwait
◯ Laos
◯ Lebanon
◯ Liberia
◯ Libya
◯ Liechtenstein
◯ Luxembourg
◯ Macedonia
◯ Madagascar
◯ Malaysia
◯ Maldives
◯ Mexico
◯ Micronesia
◯ Monaco
◯ Mongolia
◯ Morocco
◯ Nepal

◯ Netherlands
◯ New Zealand
◯ Nicaragua
◯ Nigeria
◯ North Korea
◯ Norway
◯ Pakistan
◯ Panama
◯ Papua New Guinea
◯ Paraguay
◯ Peru
◯ Philippines
◯ Poland
◯ Portugal
◯ Qatar
◯ Romania
◯ Russia
◯ Rwanda
◯ Saudi Arabia
◯ Serbia
◯ Singapore
◯ Solomon Islands
◯ Somalia
◯ South Africa
◯ South Korea
◯ Spain
◯ Sri Lanka
◯ Sudan
◯ Suriname
◯ Sweden
◯ Switzerland
◯ Taiwan
◯ Thailand
◯ Timor-Leste
◯ Tonga
◯ Turkey
◯ Ukraine
◯ United Kingdom
◯ Venezuela
◯ Vietnam
◯ Yemen
◯ Zimbabwe
◯ Other
◯ Prefer not to answer

---

Other Country: ______________________________

---

City, town, or village: ______________________________

State:

- ◯ Alabama
- ◯ Alaska
- ◯ Arizona
- ◯ Arkansas
- ◯ California
- ◯ Colorado
- ◯ Connecticut
- ◯ District of Colombia
- ◯ Delaware
- ◯ Florida
- ◯ Georgia
- ◯ Hawaii
- ◯ Idaho
- ◯ Illinois
- ◯ Indiana
- ◯ Iowa
- ◯ Kansas
- ◯ Kentucky
- ◯ Louisiana
- ◯ Maine
- ◯ Maryland
- ◯ Massachusetts
- ◯ Michigan
- ◯ Minnesota
- ◯ Mississippi
- ◯ Missouri
- ◯ Montana
- ◯ Nebraska
- ◯ Nevada
- ◯ New Hampshire
- ◯ New Jersey
- ◯ New Mexico
- ◯ New York
- ◯ North Carolina
- ◯ North Dakota
- ◯ Ohio
- ◯ Oklahoma
- ◯ Oregon
- ◯ Pennsylvania
- ◯ Rhode Island
- ◯ South Carolina
- ◯ South Dakota
- ◯ Tennessee
- ◯ Texas
- ◯ Utah
- ◯ Vermont
- ◯ Virginia
- ◯ Washington
- ◯ West Virginia
- ◯ Wisconsin
- ◯ Wyoming

State/province of residence, if any: ______________________________

Zipcode ______________________________

Zip/postal code: ______________________________

**DIAGNOSIS**

**PLEASE NOTE: Once you have completed a survey you cannot make changes to that survey. If in doubt, you can save the survey at any point and return to it once you have clarified information, e.g. with your doctor.**

**Please fill out the following for [initial_arm_1][name_patient].**

| | |
|---|---|
| Has [initial_arm_1][name_patient] been diagnosed with hydrocephalus by a medical professional? | ◯ Yes<br>◯ No<br>◯ Prefer not to answer<br>( Hydrocephalus - an abnormal accumulation of cerebrospinal (CSF) in the brain) |

**Please fill out the following for [initial_arm_1][name_patient].**

What was [initial_arm_1][name_patient]'s age when the formal diagnosis of hydrocephalus was made?

- ◯ Unknown/Unsure
- ◯ Prenatal [before birth]
- ◯ 0-3 months
- ◯ 4-7 months
- ◯ 8-11 months
- ◯ 1 year
- ◯ 2 years
- ◯ 3 years
- ◯ 4 years
- ◯ 5 years
- ◯ 6 years
- ◯ 7 years
- ◯ 8 years
- ◯ 9 years
- ◯ 10 years
- ◯ 11 years
- ◯ 12 years
- ◯ 13 years
- ◯ 14 years
- ◯ 15 years
- ◯ 16 years
- ◯ 17 years
- ◯ 18 years
- ◯ 19 years
- ◯ 20 years
- ◯ 21 years
- ◯ 22 years
- ◯ 23 years
- ◯ 24 years
- ◯ 25 years
- ◯ 26 years
- ◯ 27 years
- ◯ 28 years
- ◯ 29 years
- ◯ 30 years
- ◯ 31 years
- ◯ 32 years
- ◯ 33 years
- ◯ 34 years
- ◯ 35 years
- ◯ 36 years
- ◯ 37 years
- ◯ 38 years
- ◯ 39 years
- ◯ 40 years
- ◯ 41 years
- ◯ 42 years
- ◯ 43 years
- ◯ 44 years
- ◯ 45 years
- ◯ 46 years
- ◯ 47 years
- ◯ 48 years
- ◯ 49 years
- ◯ 50 years
- ◯ 51 years
- ◯ 52 years
- ◯ 53 years
- ◯ 54 years
- ◯ 55 years
- ◯ 56 years
- ◯ 57 years
- ◯ 58 years
- ◯ 59 years
- ◯ 60 years
- ◯ 61 years
- ◯ 62 years
- ◯ 63 years
- ◯ 64 years

◯ 65 years
◯ 66 years
◯ 67 years
◯ 68 years
◯ 69 years
◯ 70 years
◯ 71 years
◯ 72 years
◯ 73 years
◯ 74 years
◯ 75 years
◯ 76 years
◯ 77 years
◯ 78 years
◯ 79 years
◯ 80 years
◯ 81 years
◯ 82 years
◯ 83 years
◯ 84 years
◯ 85 years
◯ 86 years
◯ 87 years
◯ 88 years
◯ 89 years
◯ 90 years
◯ 91 years
◯ 92 years
◯ 93 years
◯ 94 years
◯ 95 years
◯ 96 years
◯ 97 years
◯ 98 years
◯ 99 years
◯ 100+ years

**Since [initial_arm_1][name_patient] was diagnosed before 2 years of age, we would like to know if he/she were born before his/her due date, on time, or after his/her due date.**

How many weeks away from [initial_arm_1][name_patient]'s due date was [initial_arm_1][name_patient] born?

- ◯ Unknown/Unsure
- ◯ 2 weeks late [week 42 of a pregnancy]
- ◯ 1 week late
- ◯ On time [week 40 of a pregnancy]
- ◯ 1 week premature
- ◯ 2 weeks premature
- ◯ 3 weeks premature
- ◯ 4 weeks premature
- ◯ 5 weeks premature
- ◯ 6 weeks premature
- ◯ 7 weeks premature
- ◯ 8 weeks premature
- ◯ 9 weeks premature
- ◯ 10 weeks premature
- ◯ 11 weeks premature
- ◯ 12 weeks premature
- ◯ 13 weeks premature
- ◯ 14 weeks premature
- ◯ 15 weeks premature
- ◯ 16 weeks premature
- ◯ 17 weeks premature
- ◯ 18 weeks premature
- ◯ 19 weeks premature
- ◯ 20 weeks premature [week 20 of a pregnancy]
- ◯ Prefer not to answer

**There are many ways to classify hydrocephalus. Please review the options below and select the option that most accurately describes the type of hydrocephalus that [initial_arm_1][name_patient] has. Please use the definitions to help your decision or ask your health care professional. If you do not know or are not sure, please select Unknown/Unsure.**

**Posthemorrhagic hydrocephalus of prematurity (PHH) - develops after a brain bleed in premature infants. This can be categorized as congenital or acquired, but please use this category if [initial_arm_1][name_patient] was:**
**1) Preterm AND**
**2) Had a brain bleed that led to the development of hydrocephalus**

**X-Linked Hydrocephalus (L1 Syndrome) - caused by a rare genetic abnormality in the L1CAM gene.**

**Congenital Hydrocephalus - is present at birth, but it can be diagnosed at any age (prenatal, infancy, adolescent, adulthood). When diagnosed in adults, it is often called Arrested Hydrocephalus, Compensated Hydrocephalus, or Syndrome of Hydrocephalus in Young and Middle-Aged Adults (SHYMA)**

**Acquired Hydrocephalus - caused after birth as a result of neurological conditions such as head trauma, brain tumor, cyst, intraventricular hemorrhage, or infection of the central nervous system.**

**Normal Pressure Hydrocephalus (NPH) - select NPH if the patient was diagnosed with NPH and the cause is unknown (idiopathic). NPH is usually diagnosed in older adults. Please select Acquired Hydrocephalus if the patient was diagnosed with NPH due to a known cause such as a brain infection or trauma.**

**If [initial_arm_1][name_patient] was diagnosed with**
**communicating or non-communicating hydrocephalus**
**without further specification, please ask [initial_arm_1][name_patient]'s health care professional to clarify or select unknown/unsure.**

What type of hydrocephalus does [initial_arm_1][name_patient] have?

- ◯ Unknown/Unsure
- ◯ Posthemorrhagic Hydrocephalus of Prematurity (PHH)
- ◯ X-Linked Hydrocephalus (L1 Syndrome)
- ◯ Congenital Hydrocephalus
- ◯ Acquired Hydrocephalus
- ◯ Normal Pressure Hydrocephalus (NPH) of unknown cause (idiopathic)
- ◯ Prefer not to answer

| | |
|---|---|
| What was the cause of [initial_arm_1][name_patient]'s congenital hydrocephalus? | ◯ Unknown/Unsure<br>◯ Congenital of unknown cause (idiopathic)<br>◯ Aqueductal Stenosis<br>◯ Aqueductal Pattern<br>◯ Chiari Malformation<br>◯ Dandy Walker Malformation<br>◯ Spina bifida/Myelomeningocele<br>◯ Encephalocele<br>◯ Craniosynostosis<br>◯ Arachnoid Cyst<br>◯ Posterior fossa tumor<br>◯ Supratentorial tumor<br>◯ Midbrain tumor or other midbrain lesion (e.g. pineal cyst)<br>◯ Other<br>◯ Prefer not to answer |
| If other, please specify: | __________________________________ |
| What was the cause of [initial_arm_1][name_patient]'s Acquired Hydrocephalus? | ◯ Unknown/Unsure<br>◯ Infection (e.g. meningitis, ventriculitis, encephalitis)<br>◯ Subarachnoid Hemorrhage<br>◯ Other Hemorrhage (e.g., intraventricular, Cerebellar, intracerebral)<br>◯ Head Injury, not otherwise specified<br>◯ Brain Tumor<br>◯ Other<br>◯ Prefer not to answer |
| If other, please specify: | __________________________________ |
| If you would like to provide additional detail about [initial_arm_1][name_patient]'s diagnosis, please write it here: | __________________________________ |

**Sometimes people with hydrocephalus have additional health conditions. Please let us know if [initial_arm_1][name_patient] has any of the conditions listed below.**

Does [initial_arm_1][name_patient] currently have any of the following DIAGNOSED health conditions?

(Select all that apply)

- ☐ Cerebral palsy
- ☐ Chiari malformation
- ☐ Spina bifida/myelomeningocele
- ☐ Epilepsy (multiple seizures)
- ☐ Visual Impairment
- ☐ Chronic Pain
- ☐ Migraine
- ☐ Sleep apnea [Involuntary cessation of breathing that occurs while the patient is asleep]
- ☐ Scoliosis
- ☐ Dwarfism (e.g., Achondroplasia)
- ☐ Other Growth Disorder (e.g., Human Growth Hormone (HGH) Deficiency, Intrauterine Growth Retardation (IUGR), Disproportionately Short Stature)
- ☐ Diabetes (Type 1)
- ☐ Diabetes (Type 2)
- ☐ Obesity
- ☐ Other Endocrine Disorder [disorder involving the level or combination of hormones your body produces] (e.g., Metabolic Syndrome, Hyperthyroidism, Hypothyroidism, Pituitary Disorder)
- ☐ Hyperlipidemia [An abnormally high concentration of fats or lipids in the blood]
- ☐ Hypertension [High blood pressure]
- ☐ Alzheimer's disease
- ☐ Other Dementia (e.g., Frontotemporal Dementia, Lewy Body Dementia)
- ☐ Parkinson's disease
- ☐ Other
- ☐ None of the above
- ☐ Prefer not to answer

((Select all that apply))

---

If Other Growth Disorder, please specify:

__________________________________________

---

If Other Endocrine Disorder, please specify:

__________________________________________

---

If Other Dementia, please specify:

__________________________________________

---

If Other, please specify:

__________________________________________

Has [initial_arm_1][name_patient] EXPERIENCED any of the following in the past year?

(Select all that apply.)

- ☐ Prolonged periods of pain (excluding headaches)
- ☐ Severe Pain (excluding headaches)
- ☐ Headaches
- ☐ Tiredness/Lethargy
- ☐ Shortness of Breath
- ☐ Sleep Problems
- ☐ Memory Problems
- ☐ Problems Concentrating
- ☐ Problems Navigating (e.g., trouble traveling between home and work)
- ☐ Difficulty Walking/Unable to Walk
- ☐ Dizzy/Loss of Balance
- ☐ Vision Problems
- ☐ Urinary Incontinence
- ☐ None of the above
- ☐ Prefer not to answer

((Select all that apply.))

**SURGERY**

**PLEASE NOTE: Once you have completed a survey you cannot make changes to that survey. If in doubt, you can save the survey at any point and return to it once you have clarified information, e.g. with your doctor.**

**Currently, the only effective treatments for hydrocephalus are surgical. Please answer the following questions related to Endoscopic Third Ventriculostomies (ETVs), Choroid Plexus Coagulation (CPC), and Permanent Shunts.**

**Endoscopic Third Ventriculostomy (ETV):**

| | |
|---|---|
| Has [initial_arm_1][name_patient] ever had an Endoscopic Third Ventriculostomy (ETV)? | ◯ Yes<br>◯ No<br>◯ Unknown/Unsure<br>◯ Prefer not to answer<br>(During an ETV procedure, an endoscope is used to puncture a membrane in the floor of the third ventricle creating a pathway for CSF flow within the cavities in the brain.) |
| How many ETV revision surgeries has [initial_arm_1][name_patient] undergone?<br><br>(Do not include initial ETV surgery.) | ◯ 0<br>◯ 1<br>◯ 2<br>◯ 3<br>◯ 4<br>◯ 5+<br>◯ Prefer not to answer<br>(Do not include initial ETV surgery.) |

At what age did [initial_arm_1][name_patient] have the first ETV?

(Use the date of birth as day zero.)

- ◯ Unknown/Unsure
- ◯ 0-3 months
- ◯ 4-7 months
- ◯ 8-11 months
- ◯ 1 year
- ◯ 2 years
- ◯ 3 years
- ◯ 4 years
- ◯ 5 years
- ◯ 6 years
- ◯ 7 years
- ◯ 8 years
- ◯ 9 years
- ◯ 10 years
- ◯ 11 years
- ◯ 12 years
- ◯ 13 years
- ◯ 14 years
- ◯ 15 years
- ◯ 16 years
- ◯ 17 years
- ◯ 18 years
- ◯ 19 years
- ◯ 20 years
- ◯ 21 years
- ◯ 22 years
- ◯ 23 years
- ◯ 24 years
- ◯ 25 years
- ◯ 26 years
- ◯ 27 years
- ◯ 28 years
- ◯ 29 years
- ◯ 30 years
- ◯ 31 years
- ◯ 32 years
- ◯ 33 years
- ◯ 34 years
- ◯ 35 years
- ◯ 36 years
- ◯ 37 years
- ◯ 38 years
- ◯ 39 years
- ◯ 40 years
- ◯ 41 years
- ◯ 42 years
- ◯ 43 years
- ◯ 44 years
- ◯ 45 years
- ◯ 46 years
- ◯ 47 years
- ◯ 48 years
- ◯ 49 years
- ◯ 50 years
- ◯ 51 years
- ◯ 52 years
- ◯ 53 years
- ◯ 54 years
- ◯ 55 years
- ◯ 56 years
- ◯ 57 years
- ◯ 58 years
- ◯ 59 years
- ◯ 60 years
- ◯ 61 years
- ◯ 62 years
- ◯ 63 years
- ◯ 64 years
- ◯ 65 years

◯ 66 years
◯ 67 years
◯ 68 years
◯ 69 years
◯ 70 years
◯ 71 years
◯ 72 years
◯ 73 years
◯ 74 years
◯ 75 years
◯ 76 years
◯ 77 years
◯ 78 years
◯ 79 years
◯ 80 years
◯ 81 years
◯ 82 years
◯ 83 years
◯ 84 years
◯ 85 years
◯ 86 years
◯ 87 years
◯ 88 years
◯ 89 years
◯ 90 years
◯ 91 years
◯ 92 years
◯ 93 years
◯ 94 years
◯ 95 years
◯ 96 years
◯ 97 years
◯ 98 years
◯ 99 years
◯ 100+ years
◯ Prefer not to answer
(Use the date of birth as day zero (0))

#### Choroid Plexus Coagulation (CPC):

Has [initial_arm_1][name_patient] ever had a Choroid Plexus Coagulation  (CPC)?

◯ Yes
◯ No
◯ Unknown/Unsure
◯ Prefer not to Answer
(CPC is a procedure in which a neurosurgeon uses a device to burn or cauterize tissue from the choroid plexus.  This procedure is sometimes done at the same time as an ETV (i.e., ETV/CPC,). It is not usually done on its own.)

At what age did [initial_arm_1][name_patient] first have the CPC procedure?

(Use the date of birth as day zero.)

- ◯ Unknown/Unsure
- ◯ 0-3 months
- ◯ 4-7 months
- ◯ 8-11 months
- ◯ 1 year
- ◯ 2 years
- ◯ 3 years
- ◯ 4 years
- ◯ 5 years
- ◯ 6 years
- ◯ 7 years
- ◯ 8 years
- ◯ 9 years
- ◯ 10 years
- ◯ 11 years
- ◯ 12 years
- ◯ 13 years
- ◯ 14 years
- ◯ 15 years
- ◯ 16 years
- ◯ 17 years
- ◯ 18 years
- ◯ 19 years
- ◯ 20 years
- ◯ 21 years
- ◯ 22 years
- ◯ 23 years
- ◯ 24 years
- ◯ 25 years
- ◯ 26 years
- ◯ 27 years
- ◯ 28 years
- ◯ 29 years
- ◯ 30 years
- ◯ 31 years
- ◯ 32 years
- ◯ 33 years
- ◯ 34 years
- ◯ 35 years
- ◯ 36 years
- ◯ 37 years
- ◯ 38 years
- ◯ 39 years
- ◯ 40 years
- ◯ 41 years
- ◯ 42 years
- ◯ 43 years
- ◯ 44 years
- ◯ 45 years
- ◯ 46 years
- ◯ 47 years
- ◯ 48 years
- ◯ 49 years
- ◯ 50 years
- ◯ 51 years
- ◯ 52 years
- ◯ 53 years
- ◯ 54 years
- ◯ 55 years
- ◯ 56 years
- ◯ 57 years
- ◯ 58 years
- ◯ 59 years
- ◯ 60 years
- ◯ 61 years
- ◯ 62 years
- ◯ 63 years
- ◯ 64 years
- ◯ 65 years

◯ 66 years
◯ 67 years
◯ 68 years
◯ 69 years
◯ 70 years
◯ 71 years
◯ 72 years
◯ 73 years
◯ 74 years
◯ 75 years
◯ 76 years
◯ 77 years
◯ 78 years
◯ 79 years
◯ 80 years
◯ 81 years
◯ 82 years
◯ 83 years
◯ 84 years
◯ 85 years
◯ 86 years
◯ 87 years
◯ 88 years
◯ 89 years
◯ 90 years
◯ 91 years
◯ 92 years
◯ 93 years
◯ 94 years
◯ 95 years
◯ 96 years
◯ 97 years
◯ 98 years
◯ 99 years
◯ 100+ years
◯ Prefer not to answer
(Use the date of birth as day zero (0).)

**Permanent Shunt:**

**Most Permanent Shunts are categorized into one of four (4) types:**

**VA: Ventriculo-Atrial Shunt (ventricle to heart),**
**VP: Ventriculo-Peritoneal Shunt (ventricle to abdomen),**
**VPl: Ventriculo-Pleural Shunt (ventricle to lung)**
**LP: Lumbar-Peritoneal Shunt (spine to abdomen)**

| | |
|---|---|
| Has [initial_arm_1][name_patient] ever had a Permanent Shunt? | ◯ Yes<br>◯ No<br>◯ Unknown/Unsure<br>◯ Prefer not to answer<br>((i.e., VP, VA, VPl, LP)) |
| Does [initial_arm_1][name_patient] have a permanent shunt now? | ◯ Yes<br>◯ No<br>◯ Unknown/Unsure<br>◯ Prefer not to answer<br>((i.e., VA, VP, VPl, LP)) |
| How many permanent shunts does [initial_arm_1][name_patient] CURRENTLY | ◯ One (1) permanent shunt<br>◯ More than 1 permanent shunt<br>◯ Unknown/Unsure<br>◯ Prefer not to answer<br>(1. One permanent proximal catheter located in the ventricle (VP, VA, or VPl) or One lumbar-peritoneal (LP) shunt.<br>2. More than one permanent proximal catheter located in the ventricle(s) or At least one ventriculo- (VP, VA, or VPl) and one lumbar-peritoneal (LP) shunt.) |
| What type(s) of shunt(s) does [initial_arm_1][name_patient] have?<br><br>(Select all that apply) | ☐ Ventricular-Atrial Shunt (VA, ventricle to heart)<br>☐ Ventricular-Peritoneal Shunt (VP, ventricle to abdomen)<br>☐ Ventricular-Pleural Shunt (VPl, ventricle to lung)<br>☐ Lumbar-Peritoneal Shunt (LP, spine to abdomen)<br>☐ Other<br>☐ Unknown/Unsure<br>☐ Prefer not to answer<br>((Select all that apply) ) |

At what age did [initial_arm_1][name_patient] have the first shunt surgery?

(Use the date of birth as day zero.)

◯ Unknown/Unsure
◯ 0-3 months
◯ 4-7 months
◯ 8-11 months
◯ 1 year
◯ 2 years
◯ 3 years
◯ 4 years
◯ 5 years
◯ 6 years
◯ 7 years
◯ 8 years
◯ 9 years
◯ 10 years
◯ 11 years
◯ 12 years
◯ 13 years
◯ 14 years
◯ 15 years
◯ 16 years
◯ 17 years
◯ 18 years
◯ 19 years
◯ 20 years
◯ 21 years
◯ 22 years
◯ 23 years
◯ 24 years
◯ 25 years
◯ 26 years
◯ 27 years
◯ 28 years
◯ 29 years
◯ 30 years
◯ 31 years
◯ 32 years
◯ 33 years
◯ 34 years
◯ 35 years
◯ 36 years
◯ 37 years
◯ 38 years
◯ 39 years
◯ 40 years
◯ 41 years
◯ 42 years
◯ 43 years
◯ 44 years
◯ 45 years
◯ 46 years
◯ 47 years
◯ 48 years
◯ 49 years
◯ 50 years
◯ 51 years
◯ 52 years
◯ 53 years
◯ 54 years
◯ 55 years
◯ 56 years
◯ 57 years
◯ 58 years
◯ 59 years
◯ 60 years
◯ 61 years
◯ 62 years
◯ 63 years
◯ 64 years
◯ 65 years

| | |
|---|---|
| | ◯ 66 years<br>◯ 67 years<br>◯ 68 years<br>◯ 69 years<br>◯ 70 years<br>◯ 71 years<br>◯ 72 years<br>◯ 73 years<br>◯ 74 years<br>◯ 75 years<br>◯ 76 years<br>◯ 77 years<br>◯ 78 years<br>◯ 79 years<br>◯ 80 years<br>◯ 81 years<br>◯ 82 years<br>◯ 83 years<br>◯ 84 years<br>◯ 85 years<br>◯ 86 years<br>◯ 87 years<br>◯ 88 years<br>◯ 89 years<br>◯ 90 years<br>◯ 91 years<br>◯ 92 years<br>◯ 93 years<br>◯ 94 years<br>◯ 95 years<br>◯ 96 years<br>◯ 97 years<br>◯ 98 years<br>◯ 99 years<br>◯ 100+ years<br>◯ Prefer not to answer<br>(Use the date of birth as day zero (0).) |
| Does [initial_arm_1][name_patient]'s shunt have a programmable valve? | ◯ Yes<br>◯ No<br>◯ Unsure/Unknown<br>◯ Prefer not to answer<br>(If the patient has multiple shunts, select Yes if any are programmable.) |

How many shunt revisions has [initial_arm_1][name_patient] had?

(Do not include the initial placement)

- ◯ Unknown/Unsure
- ◯ 0
- ◯ 1
- ◯ 2
- ◯ 3
- ◯ 4
- ◯ 5
- ◯ 6
- ◯ 7
- ◯ 8
- ◯ 9
- ◯ 10
- ◯ 11
- ◯ 12
- ◯ 13
- ◯ 14
- ◯ 15
- ◯ 16
- ◯ 17
- ◯ 18
- ◯ 19
- ◯ 20-29
- ◯ 30-39
- ◯ 40-49
- ◯ 50-59
- ◯ 60-69
- ◯ 70-79
- ◯ 80-89
- ◯ 90-99
- ◯ 100-149
- ◯ 150-199
- ◯ 200+
- ◯ Prefer not to answer

(Do not include initial shunt placement. Only include surgeries during which a part or all of the shunt was replaced.)

How many shunt infections has [initial_arm_1][name_patient] had?

- ◯ Unknown/Unsure
- ◯ 0
- ◯ 1
- ◯ 2
- ◯ 3
- ◯ 4
- ◯ 5
- ◯ 6
- ◯ 7
- ◯ 8
- ◯ 9
- ◯ 10
- ◯ 11
- ◯ 12
- ◯ 13
- ◯ 14
- ◯ 15
- ◯ 16
- ◯ 17
- ◯ 18
- ◯ 19
- ◯ 20-29
- ◯ 30-39
- ◯ 40-49
- ◯ 50+
- ◯ Prefer not to answer

**MEDICAL CARE AND INSURANCE**

**PLEASE NOTE: Once you have completed a survey you cannot make changes to that survey. If in doubt, you can save the survey at any point and return to it once you have clarified information, e.g. with your doctor.**

**We would like to know what types of doctors [initial_arm_1][name_patient] goes to and what types of insurance [initial_arm_1][name_patient] has. Please answer the following questions related to medical care and insurance.**

**Medical Care Team:**

**Primary Care Physician:**

Who is [initial_arm_1][name_patient]'s primary care physician (PCP)?

- ◯ Does not have one
- ◯ Pediatrician
- ◯ Family Practice Doctor
- ◯ Advance Practice Provider (e.g. Nurse Practitioner, Clinical Nurse Specialist)
- ◯ Internist
- ◯ OB/GYN
- ◯ Gerontologist
- ◯ Other
- ◯ Unknown/Unsure
- ◯ Prefer not to answer

(Do not include Neurosurgeons or Neurologists.)

#### Neurosurgeon:

| | |
|---|---|
| Does [initial_arm_1][name_patient] have a neurosurgeon? | ◯ Yes<br>◯ No<br>◯ Unknown/Unsure<br>◯ Prefer not to answer |
| How often does [initial_arm_1][name_patient] see the neurosurgeon for routine checkups? | ◯ More than once a year<br>◯ Once a year<br>◯ Every 2-3 years<br>◯ Every 4 years or more<br>◯ Does not see him/her for routine checkups (only sees him/her when there are problems)<br>◯ Unknown/unsure<br>◯ Prefer not to answer |

#### Neurologist:

| | |
|---|---|
| Does [initial_arm_1][name_patient] have a neurologist? | ◯ Yes<br>◯ No<br>◯ Unknown/Unsure<br>◯ Prefer not to answer |
| How often does [initial_arm_1][name_patient] see the neurologist for routine checkups? | ◯ More than once a year<br>◯ Once a year<br>◯ Every 2-3 years<br>◯ Every 4 years or more<br>◯ Does not see him/her for routine checkups (only sees him/her when there are problems)<br>◯ Unknown/unsure<br>◯ Prefer not to answer |

#### Insurance:

What type of health insurance/health care plan, if any, does [initial_arm_1][name_patient] currently have?

(Select all that apply.)

- ☐ Unknown/Unsure
- ☐ No Coverage
- ☐ Has Coverage but unsure of type
- ☐ Private health insurance (including ACA/Obamacare)
- ☐ Medicare
- ☐ Medi-gap
- ☐ Medicaid
- ☐ CHIP (Children's Health Insurance Program)
- ☐ Military health care (Tricare/VA, Champ-VA)
- ☐ Indian health service
- ☐ State-sponsored health plan
- ☐ Other government program
- ☐ Single service plan (e.g., dental, vision, prescription)
- ☐ Prefer not to answer

((Select all that apply.) )

---

What is the source of [initial_arm_1][name_patient]'s primary health insurance?

- ◯ Unknown/Unsure
- ◯ Employer/former employer (including military) insurance plan
- ◯ His/Her own private insurance plan
- ◯ His/Her own government-sponsored insurance plan (e.g., Medicare, Medicaid)
- ◯ Parent's insurance plan
- ◯ Spouse's/other relative's insurance plan
- ◯ Other
- ◯ Prefer not to answer

---

Has [initial_arm_1][name_patient] ever had difficulty receiving any of the following because of high out of pocket cost?

(Select all that apply.)

- ☐ Unknown/Unsure
- ☐ Operations/Procedures
- ☐ Access to Medical specialists
- ☐ Physical/Occupational/Speech-language therapy, rehabilitation services
- ☐ Psychological counseling
- ☐ Neuropsychological testing
- ☐ Medications
- ☐ Genetic testing
- ☐ Assistive devices/Orthotics (wheelchair, braces, helmet)
- ☐ Other
- ☐ No difficulty receiving any of the above
- ☐ Prefer not to answer

((Select all that apply.) )

---

If other, please specify

__________________________________

---

In non-emergency situations, when [initial_arm_1][name_patient] or you suspects there may be a hydrocephalus-related problem, whom does [initial_arm_1][name_patient] or you contact first?

(e.g., a shunt/ETV problem)

- ◯ Primary care physician - by choice
- ◯ Primary care physician - because a referral is required
- ◯ Specialized clinic (e.g., spina bifida clinic)
- ◯ Neurologist
- ◯ Neurosurgeon
- ◯ Emergency room
- ◯ Other
- ◯ Unknown/Unsure
- ◯ Prefer not to answer

**Learning, Cognitive, and Movement Disorders**

**PLEASE NOTE: Once you have completed a survey you cannot make changes to that survey. If in doubt, you can save the survey at any point and return to it once you have clarified information, e.g. with your doctor.**

Has [initial_arm_1][name_patient] ever been DIAGNOSED with any of the following educational disorders?

(Select all that apply.)

- ☐ Learning disability in math
- ☐ Learning disability in reading
- ☐ Writing disability
- ☐ Non-verbal learning disability
- ☐ None of the above
- ☐ Unknown/Unsure
- ☐ Prefer not to answer

((Select all that apply.) )

Has [initial_arm_1][name_patient] ever been DIAGNOSED with any of the following cognitive disorders?

(Select all that apply.)

- ☐ Attention Deficit Hyperactivity Disorder (ADHD, including hyperactive or inattentive or both)
- ☐ Autism Spectrum Disorder (includes pervasive development disorder, Asperger's, high functioning autism)
- ☐ Language (receptive) delay (problems understanding language)
- ☐ Speech (expressive) delay (problems getting words out)
- ☐ Executive Function Disorder
- ☐ Global Developmental Delay
- ☐ Intellectual Disability
- ☐ Visual Processing Deficits
- ☐ Short term memory problems
- ☐ None of the above
- ☐ Unknown/Unsure
- ☐ Prefer not to answer

((Select all that apply.) )

Has [initial_arm_1][name_patient] ever been DIAGNOSED with any of the following emotional disorders?

(Select all that apply.)

- ☐ Depression
- ☐ Anxiety
- ☐ Obsessive Compulsive Disorder
- ☐ None of the above
- ☐ Unknown/Unsure
- ☐ Prefer not to answer

((Select all that apply.) )

Has [initial_arm_1][name_patient] ever EXPERIENCED any of the following issues with movement?

(Select all that apply.)

- ☐ Clumsiness (Dyspraxia)
- ☐ Tremors
- ☐ Tics
- ☐ Other stereotyped movements [Stereotyped movements are persistent and repetitive]
- ☐ Trouble with balance
- ☐ Trouble with walking/Unable to walk
- ☐ None of the above
- ☐ Unknown/Unsure
- ☐ Prefer not to answer

((Select all that apply.) )

PLEASE NOTE: Once you have completed a survey you cannot make changes to that survey.
If in doubt, you can save the survey at any point and return to it once you have clarified information, e.g. with your doctor.
